# A Systematic Review and Meta-Analysis on Mental Illness Symptoms in Spain in the COVID-19 Crisis

**DOI:** 10.1101/2021.04.11.21255274

**Authors:** Richard Z. Chen, Stephen X. Zhang, Wen Xu, Allen Yin, Rebecca Kechen Dong, Bryan Z. Chen, Andrew Delios, Roger S. McIntyre, Saylor Miller, Xue Wan

## Abstract

**Objective:** This paper systematically reviews and assesses the prevalence of anxiety, depression, and insomnia symptoms in the general population, frontline healthcare workers (HCWs), and adult students in Spain during the COVID-19 crisis.

**Data sources:** Articles in PubMed, Embase, Web of Science, PsycINFO, and medRxiv from March 2020 to February 6, 2021.

**Results:** The pooled prevalence of anxiety symptoms in 23 studies comprising a total sample of 85,560 was 20% (95% CI: 15% - 25%, I2 = 99.9%), that of depression symptoms in 23 articles with a total sample comprising of 86,469 individuals was 23% (95% CI: 18% - 28%, I2 = 99.8%), and that of insomnia symptoms in 4 articles with a total sample of 915 were 52% (95% CI: 42-64%, I2 = 88.9%). The overall prevalence of mental illness symptoms in frontline HCWs, general population, and students in Spain are 42%, 19%, and 50%, respectively.

**Discussion:** The accumulative evidence from the meta-analysis reveals that adults in Spain suffered higher prevalence rates of mental illness symptoms during the COVID-19 crisis with a significantly higher rate relative to other countries such as China. Our synthesis reveals high heterogeneity, varying prevalence rates and a relative lack of studies in frontline and general HCWs in Spain, calling future research and interventions to pay attention to those gaps to help inform evidence-based mental health policymaking and practice in Spain during the continuing COVID-19 crisis. The high prevalence rates call for preventative and prioritization measures of the mental illness symptoms during the Covid-19 pandemic.

## 1. Introduction

Since its first report on November 17, 2019, the COVID-19 pandemic has resulted in more than 130 million confirmed cases and almost 3 million deaths. Spain, with its first case on January 31, 2020, is one of the most affected countries in Europe by COVID-19 infections, complications, and deaths. The COVID-19 crisis in Spain is particularly severe as a quarter of the population have been unable to isolate themselves according to public health recommendations^3^. Like most parts of the world, the COVID-19 pandemic has severely affected the Spanish population and has significantly altered the workplace and day-to-day activity. Disruptions such as social distancing laws, mandatory lockdowns, and a high risk of infection can lead to psychological problems^4^. A survey study of 17 countries by Luo et al.^5^ found that Spain had one of the highest prevalence rates of anxiety during the Covid-19 pandemic.

Rising rates of mental illness as well as the correlation to reduced mental health quality have been reported globally with differences across regions^6^. For example, it has been reported that the prevalence of mild anxiety symptoms in Spain was variably reported from 1.8% ^2^ - 58.6%^7^ while mild depression symptoms have varied from 7.3% ^2^ - 46.0%^7^. Given the disparity, it is pertinent to implement a meta-analysis to pool empirical study prevalence rates to accumulate meta-analytical evidence. Such meta-analytical evidence on the prevalence rate of mental health issues in Spain can help with evidence-based decisions on the allocation of limited resource during the continuing COVID-19 crisis.

This study aims to conduct the first systematic review and meta-analyses on the mental illness symptoms of the Spain population by assessing the prevalence of anxiety, depression, and insomnia symptoms during the COVID-19 virus. Our primary focus was on populations of healthcare workers, general adult population, and students as the foregoing populations have been reported by others to be at greater risk of negative mental health outcomes. ^8^

## 2. Methods and Materials

We registered with the International Prospective Register of Systematic Reviews (PROSPERO: CRD42020224458) and used the Preferred Reporting Items for Systematic Reviews and Meta-Analyses (PRISMA) statement 2019 to guide our search.

### 2.1 Data Sources and Search Strategy

This paper comes from a large project on meta-analysis of mental illness symptoms during COVID-19. We searched the following databases for studies that fit our criteria: *PubMed, Embase*, PsycINFO, and *Web of Science* from Feb 2020 to Feb 6^th^, 2021. To identify articles based off the requirements, we searched specific titles and abstracts using keywords in Table S1 with Boolean operators. We also searched the preprints at medRxiv.

### 2.2 Selection Criteria

The search included empirical studies that reported the prevalence of anxiety, depression, or insomnia symptoms among frontline HCWs, general HCWS, general adult populations, or adult (university) students in Spain. To be included, reports had to implement validated psychometric measures with outcomes reported in English.

We excluded studies that had populations of children, adolescents, or adult subpopulations (e.g., pregnant women). We also excluded non-original research or studies which were reviews, meta-analyses, qualitative and case studies, interviews, news reports, interventional studies, or articles without validated instruments or validated cutoff scores to identify prevalence.

A researcher (WX) contacted the authors of papers that missed important information in several instances: 1) if they surveyed a population that included both targeted and excluded populations in a way that we could not identify the prevalence rate for our desired population; 2) if the paper included primary data meeting our inclusion criteria, but did not report the prevalence; 3) if the paper reported the overall prevalence without specifying whether it is mild above or moderate above: or 4) if the paper was missing or unclear about critical information such as respondent rate, data collection time, or female proportion rate.

### 2.3 Data Screening

To begin, we exported article information from various databases into Endnotes to remove duplicates and then imported them into Rayyan. Two researchers independently (BZC & AD) screened the titles and abstracts of all papers using our inclusion and exclusion criteria. Any conflicts were resolved by a third researcher (RZD).

### 2.4 Data Extraction

A well-developed coding protocol and coding book were developed based on previous studies ^9^. All included articles from the screening were sent to two pairs of researchers who were assigned to thoroughly examine and extract important data into a coding book (WX & AY, BZC & AD, RZC & SM). Using a coding procedure, the coders coded relevant information including author, title, country, starting and ending dates of data collection, study design, population, sample size, respondent rate, female proportion rate, age range and mean, outcome, outcome level, instruments, cutoff scores, and prevalence.

Comments for the reason behind emailing and excluding papers were recorded. After both coders had independently coded their articles, they would then crosscheck their information with one another. Different answers were conferred with and changed, and disagreements were settled through a third coder to code the article and settle the disagreement. The lead coder (RZC) double-checked important data including the population, sample size, mental health outcomes, outcome levels, instruments, and prevalence. Papers with unusual prevalence, cutoff scores, and numbers were double-checked afterwards for sensitivity analysis.

### 2.5 Assessment of Bias Risk

The Mixed Methods Appraisal Tool (MMAT)^10^, a seven question test that assesses the quality of research papers was used for our meta-analyses. Pairs of coders individually coded these questions. Discrepancies were once resolved through the lead coder (RZD). The quality scores of papers were on a range from 0 to 7. Studies with a quality of above 6 were considered high, articles with a score of between 5 and 6 were classed as medium, and articles with a score of below 5 were considered low.

### 2.6 Data analysis

Using Version 16.1 of Stata, we conducted a random effect meta-analysis to calculate the pooled prevalence from multiple studies using meta-prop. We used the *I*^*2*^ statistics to examine the heterogeneity of prevalence among studies and the heterogeneity was classed as high when *I*^*2*^ is higher than 75%^11^.

## 3 Results

### 3.1 Study Screening

Figure 1 illustrates the PRISMA flow chart on our method of search and data extraction process. We found 6949 citations from the selected database and other sources. After excluding 2729 entries that did not meet the including criteria in the screening process, we extracted data from 684 citations based on its full text. After excluding 505 citations in the data extraction process, we coded 150 papers that included the necessary information to conduct meta-analyses. In addition, we sent out two rounds of emails to the authors of 95 papers to request useful information. Among them, we have received 75 responses. Among these responses, we received new prevalence data from 8 out of the 29 studies that lacked prevalence data. Finally, we had 168 studies providing necessary information for meta-analysis, among which, 30 articles had participants that were from Spain.

**Figure 1.**
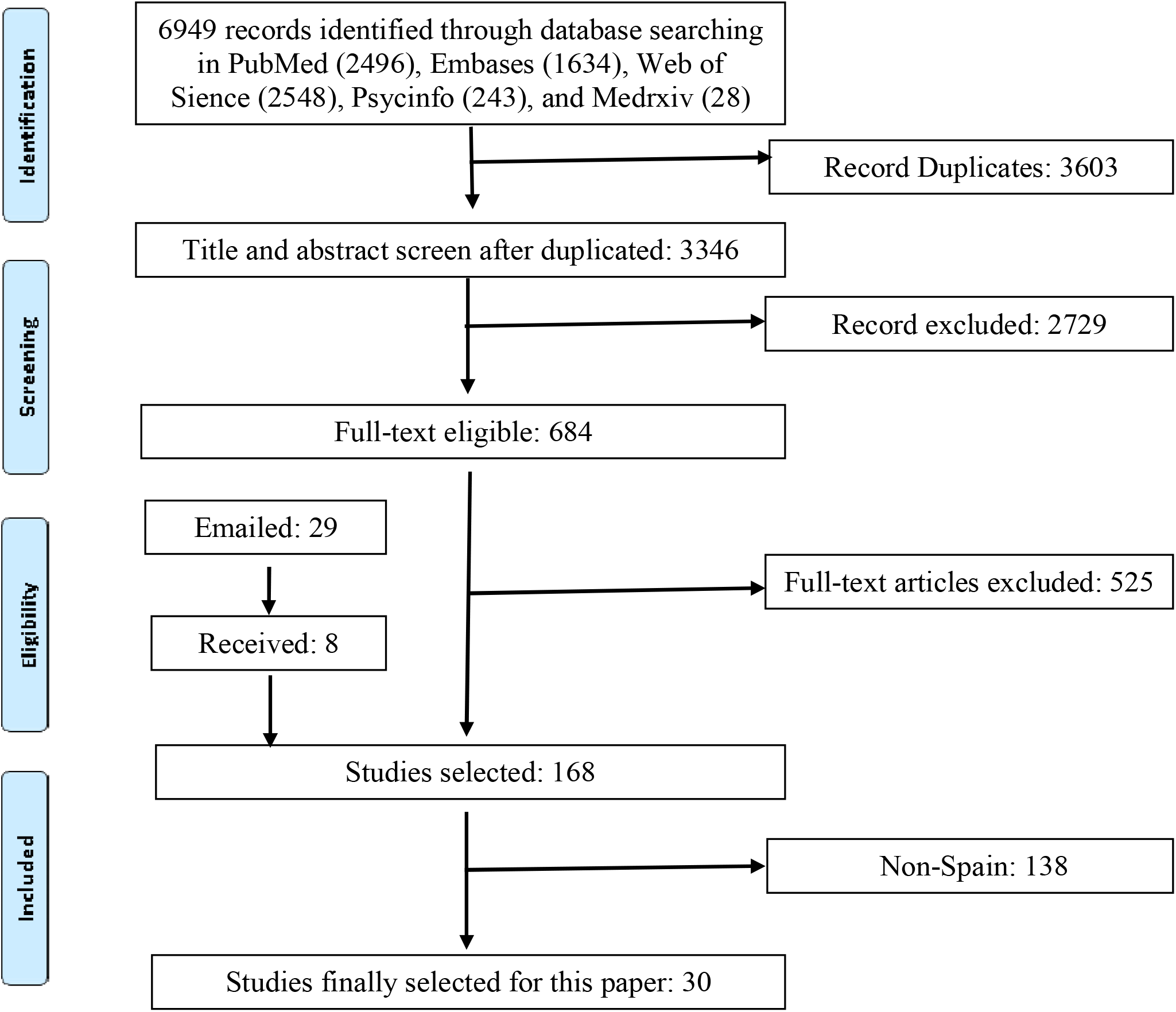
PRISMA flow diagram.

### 3.2 Study Characteristics

In total, 41 samples from 30 studies ^7,12-39^ were included in this meta-analysis with 90,036 participants from Spain. Among them, 32 samples (78.1%) were of general populations, 4 samples of frontline HCW’s (9.8%), and 5 (122%) of adult students. Most studies were cross-sectional (86.7%) and 4 studies were longitudinal cohort studies study (13.3%). Almost all studies were published (93.2%) while only 2 (6.7%) remained as preprints. The sample size of the 41 samples varies from 44 to 21207 with its medium value of 1174. The participation rates varied from 20.0% to 98.0% with the medium of 70.3%. The female proportions among the 41 samples varied from 0% to 100% with the medium value of 86.7%.

### 3.3 The pooled prevalence of anxiety, depression, and insomnia

In Spain, 30 samples from 23 studies reported the prevalence of anxiety symptoms among 85,560 participants. Several anxiety instruments were used, including the Depression, Anxiety and Stress Scale - 21 Items (DASS-21) being used the most (56.5%), followed by Generalized Anxiety Symptoms 7-items scale (GAD-7) (30.4%), Hospital Anxiety and Depression Scale (HADS) (4.4%), and Brief Symptom Inventory 18 (BSI-18) (4.4%). Different studies used different cut-off values to determine the overall prevalence as well as the severity of anxiety. In the random-effects model, the pooled prevalence of anxiety was 20% (95% CI: 15% - 25%, I2 = 99.9%) in the 23 studies (Figure 2A).

**Figure 2A.**
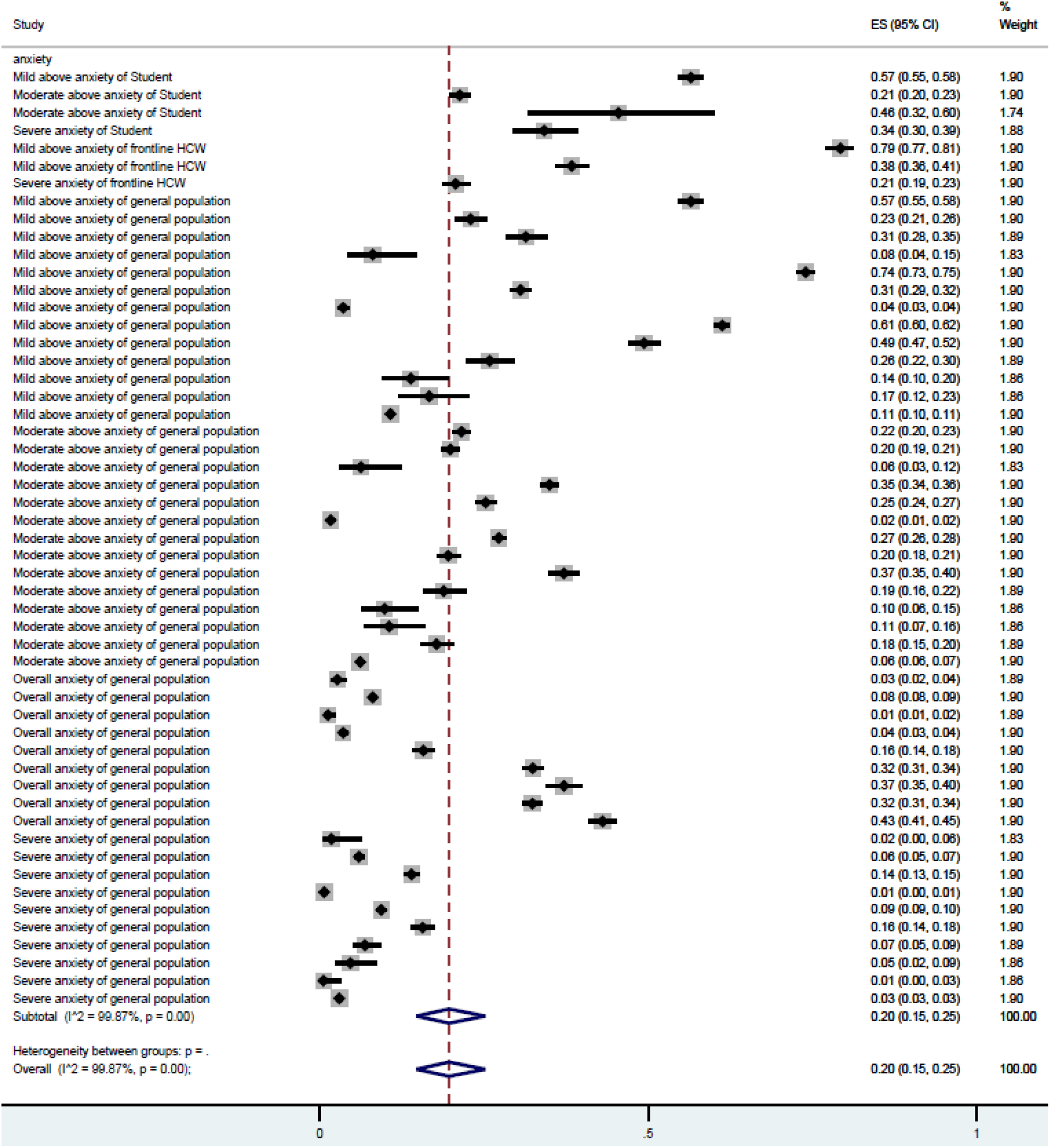
Forest plot of the prevalence of anxiety. Figure legend: The square markets indicate the prevalence of anxiety at the different level for different population. The size of the marker correlates to the inverse variance of the effect estimate and indicates the weight of the study. The diamond data market indicates the pooled prevalence.

A total of 31 samples of our total 23 articles that we reported in this meta-analysis were on depression, for a total of 86,649 respondents. Several depression instruments were used including DASS-21 being used the most (52.2%), followed by Patient Health Questionnaire (PHQ)-9 (30.4%), Beck Depression Inventory (BDI) (4.4%), HADS (4.4%), and BSI (4.4%). In the random-effects model, the pooled prevalence of depression was 23% (95% CI: 18% - 28%, I2 = 99.8%) among the 23 studies (Figure 2B).

**Figure 2B.**
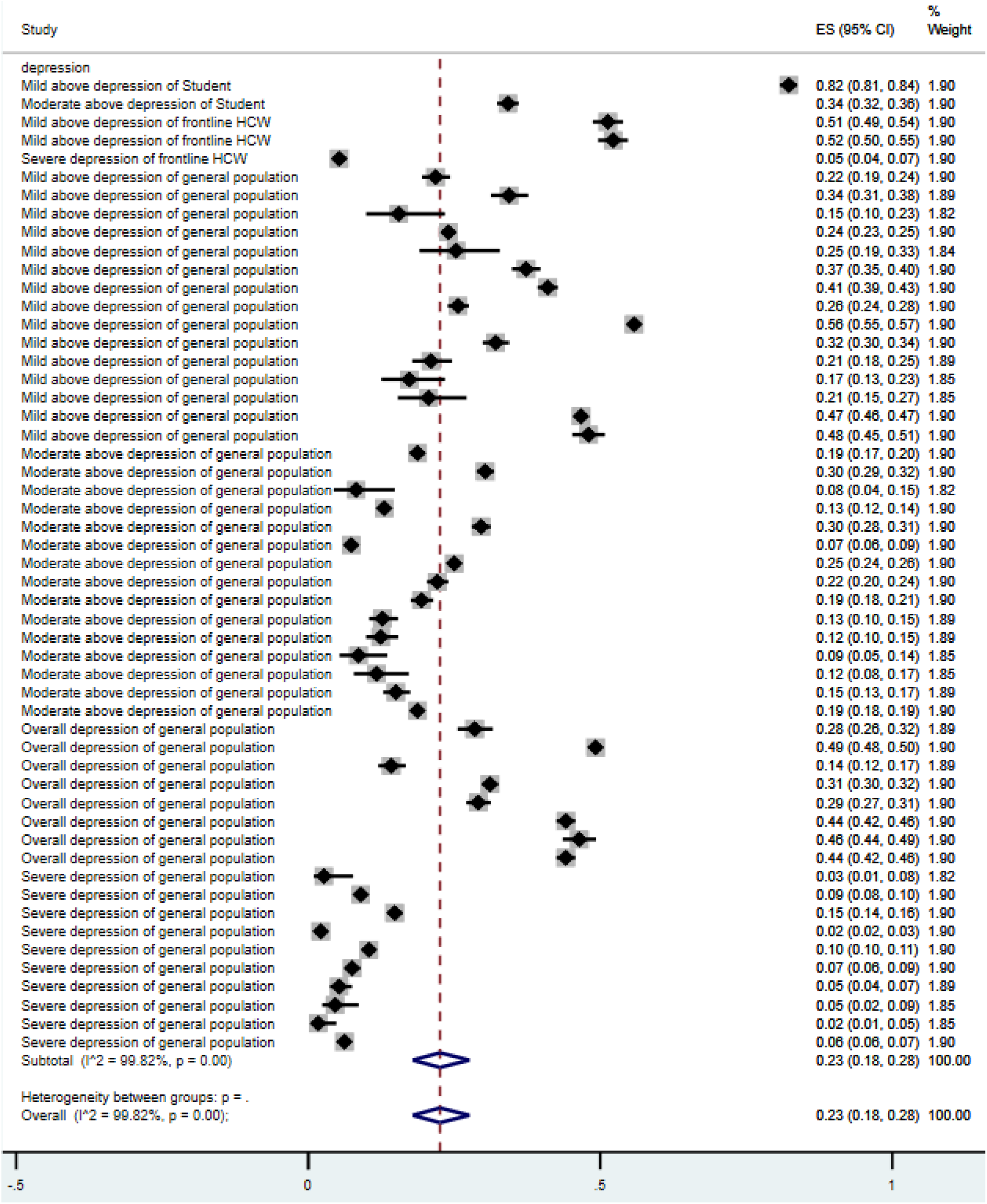
Forest plot of the prevalence of depression. Figure legend: The square markets indicate the prevalence of anxiety at the different level for different population. The size of the marker correlates to the inverse variance of the effect estimate and indicates the weight of the study. The diamond data market indicates the pooled prevalence.

Seven samples of the 4 articles that we reported in this meta-analysis reported on insomnia, for a total of 915 respondents. The Insomnia Severity Index (ISI) and Pittsburgh Sleep Quality Index (PSQI) were used to measure insomnia. In the random-effects model, the pooled prevalence of insomnia is 52% (95% CI: 42-64%, I2 = 88.9%) (Figure 2C).

**Figure 2C.**
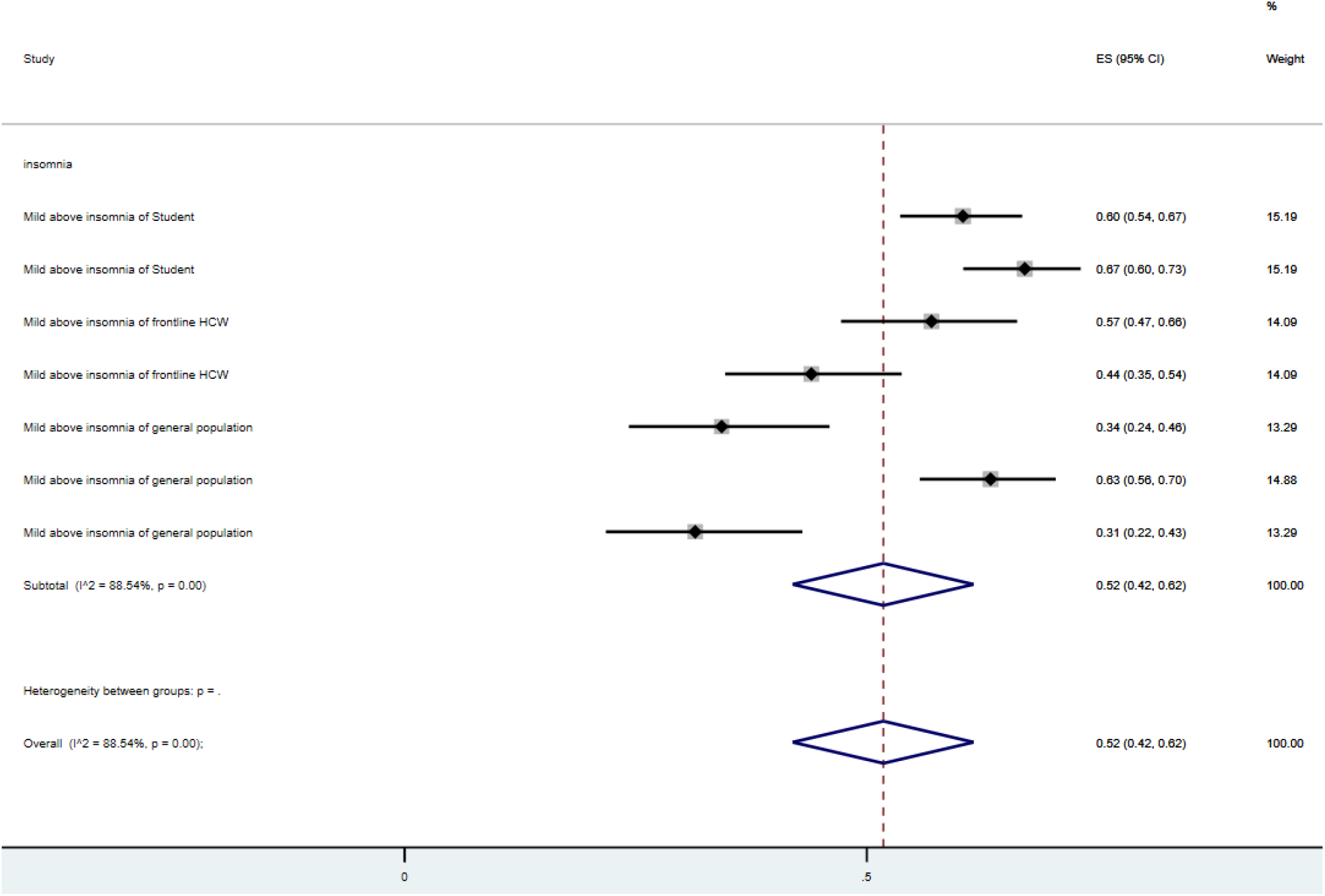
Forest plot of the prevalence of insomnia. Figure legend: The square markets indicate the prevalence of anxiety at the different level for different population. The size of the marker correlates to the inverse variance of the effect estimate and indicates the weight of the study. The diamond data market indicates the pooled prevalence.

The overall prevalence of mental health disorder in frontline students HCWs, and general population in Spain are 50%, 42%, and 19% respectively. The overall prevalence rates of mental illness symptoms that surpassed the cutoff values of mild, moderate, and severe symptoms were 38%, 18%, and 7%, respectively.

### 3.5 Quality of articles

Using the Mixed Methods Appraisal Tool (MMAT)^18^, we found that 7 studies (22.3%) are of higher quality, 23 studies (76.7%) had a medium quality, and 0 studies were of low quality (Table 1). The subgroup analysis suggests the studies with high quality reported lower prevalence of mental health issues in Spain (Table 2).

**Table 1.** Study Characteristics on mental illness symptoms in COVID-19 epidemic in Spain.

| Characteristics | Total number of studies/samples* | Percent (%) | Level of analysis |
| --- | --- | --- | --- |
| *Overall* | 30/41 | 100 |  |
| *Design* |  |  | Study |
| Cohort | 4 | 13.3 |  |
| Cross-sectional | 26 | 86.7 |  |
| *Publication status* |  |  | Study |
| Preprint | 2 | 6.7 |  |
| Published | 28 | 93.2 |  |
| *Quality* |  |  | Study |
| >6 | 7 | 23.3 |  |
| Between 5 and 6 | 23 | 76.7 |  |
| <5 | 0 | 0.0 |  |
| *Population* |  |  | Sample |
| Frontline HCW | 4 | 9.8 |  |
| General population | 32 | 78.1 |  |
| Student | 5 | 12.2 |  |
| *Outcome#* |  |  | Sample |
| Anxiety | 53 | 46.9 |  |
| Depression | 53 | 46.9 |  |
| Insomnia | 7 | 6.2 |  |
| *Severity#* |  |  | Sample |
| Above mild | 41 | 36.3 |  |
| Above moderate | 32 | 27.3 |  |
| Severe | 23 | 29.4 |  |
| Overall | 17 | 15.0 |  |
|  | Median (mean) | Range |  |
| *Sample size* | 1174 (2196) | 44 - 21207 | Sample |
| *Response rate* | 70.3% (75.4%) | 20.0% - 98.0% | Sample |
| *Female portion* | 86.7% (68.5%) | 0% -100% | Sample |

\* One study may include multiple independent samples such as frontline HCW and general population.

**Table 2.** The pooled prevalence rates of mental illness symptoms by subgroups of population, outcome, and severity.

| First-level subgroup | Second-level subgroup | Prevalence (%) | 95% CI | I<sup>2</sup> (%) | P value |
| --- | --- | --- | --- | --- | --- |
| *Overall* |  | 22% | 19% - 25% | 99.84% | 0.00 |
| *Population* | Frontline HCW | 42% | 23% - 63% | 99.73% | 0.00 |
|  | General population | 19% | 16% - 23% | 99.85% | 0.00 |
|  | Student | 50% | 32% - 69% | 99.73% | 0.00 |
| *Outcome#* | Anxiety | 20% | 15% - 24% | 99.87% | 0.00 |
|  | Depression | 23% | 18% - 28% | 99.82% | 0.00 |
|  | Insomnia | 52% | 42% - 62% | 88.54% | 0.00 |
| *Severity#* | Above mild | 38% | 31% - 45% | 99.82% | 0.00 |
|  | Above moderate | 18% | 15% - 22% | 99.54% | 0.00 |
|  | Severe | 7% | 5% - 9% | 99.05% | 0.00 |
|  | Overall | 25% | 16% - 34% | 99.83% | 0.00 |
| *Quality* | Studies with high quality | 21% | 15% - 27%, | 99.0% | 0.00 |
|  | Studies with medium quality | 23% | 19% -27% | 99.9% | 0.00 |

Note: CI = Confidence Interval; I<sup>2</sup> statistic indicates the heterogeneity.

### 3.6 Sensitivity Analysis

Given this study pooled the proportion studies in a meta-analysis and research methodologists have found the conventional funnel plots to assess biases in meta-analyses are inaccurate for meta-analyses of proportion studies^40^. In meta-analyses of proportion study, a Doi plot and the Luis Furuya–Kanamori (LFK) index represent the better approach for graphically representing publication bias – where a symmetrical triangle implies the absence of publication bias, while an asymmetrical triangle indicates possible publication bias. ^41^ The Doi plot and LFK index have higher sensitivity and power to detect publication bias than the funnel plot and Egger’s regression^42^.The LFK index provides a quantitative measure to assess the of asymmetry - a score within ±1 indicates ‘no asymmetry’, exceeds ±1 but is within ±2 indicates ‘minor asymmetry’ and exceeds ±2 indicates ‘major asymmetry’. Figure 3 depicts the Doi plot and a Luis Furuya–Kanamori (LFK) index 0f −0.45, indicating ‘no asymmetry’ and the unlikely presence of publication bias. We also tested the impact of publication status and sample size and did not find significant influence.

**Figure 3.**
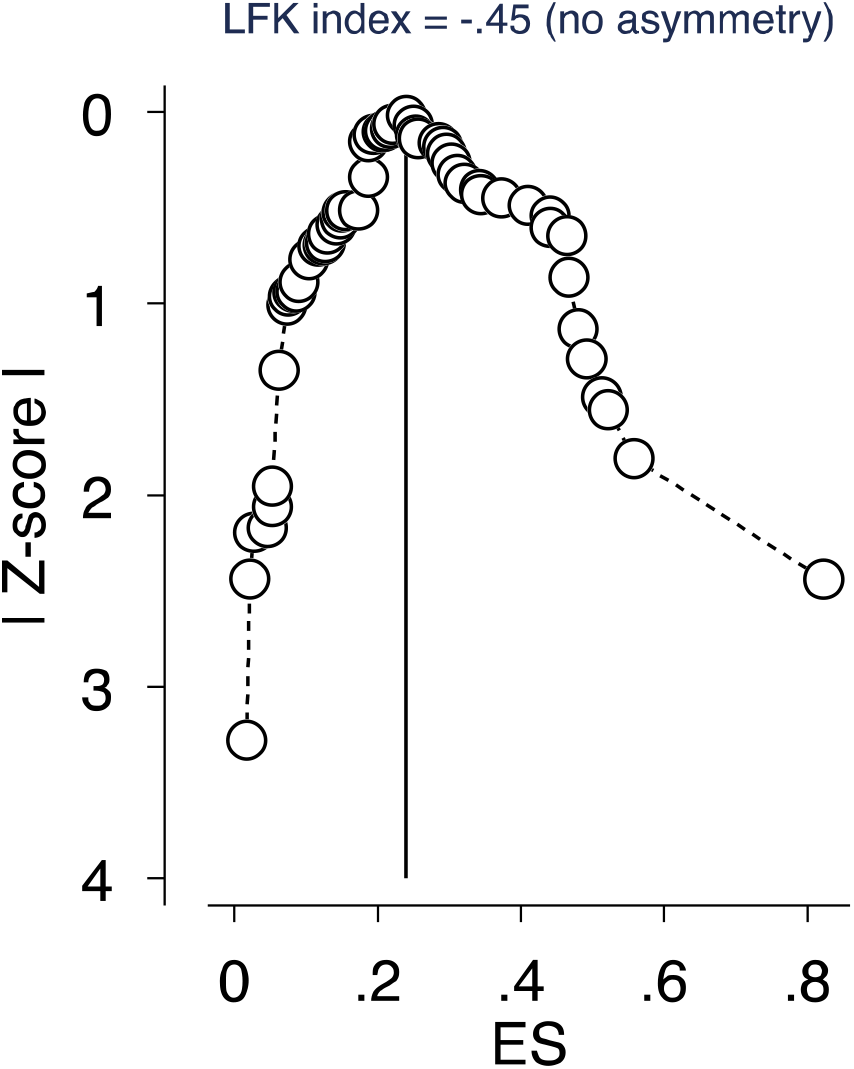
Depiction of publication bias in the baseline meta-analysis of proportion studies based on Doi plot and the Luis Furuya–Kanamori (LFK) index -a score that is within ±1 indicates ‘no asymmetry’

## 4 Discussion

### 4.1 Overview of the findings

To date, this meta-analysis is the first to report the pooled prevalence of the mental illness symptoms on key populations in Spain during the COVID-19 crisis. Our study included data from a total of 41 samples and 30 articles for a total of 90,036 adults in a single year of the COVID-19 pandemic. The pooled prevalence rates of anxiety, depression, and insomnia symptoms at the mild above level are 34%, 36%, and 52% respectively (Appendix 2). The analysis found that both students and frontline HCWs had much higher prevalence rates of mental illness than the general population did. The foregoing finding of greater risk in students and frontline HCWs is in accordance with what has been reported elsewhere. The higher prevalence in students may be due to their tendencies to worry over the health of both themselves and loved ones as well as the pressures they face ^43 44^, and the daily interactions and jobs under high-risk virus transmission, for the frontline HCWs ^45^.

Insomnia in Spain appeared to be much more prevalent than anxiety and depression with 52% of the population being diagnosed with insomnia. Part of the reason for this might be that all the 7 samples on the prevalence of insomnia used a mild above severity level, yielding a higher number prevalence rate.

### 4.2 Comparison with Prior Meta-analyses

Given that this meta-analysis is the first on the topic in Spain, it is worthwhile to compare the findings with other similar meta-analyses in other countries. The existing meta-analyses on COVID-19 mental health were conducted mostly on a few geographical regions, with the majority of them being done in China, the first country to experience the COVID-19 crisis. The pooled prevalence of anxiety symptoms in Spain population was 34%, which is much higher than, for example reported in Bareeqa et al. (2020) (22%), Pappa et al. (2020) (23%), Krishnamoorthy et al. 2020 (26%) and Ren et al. 2020 (25%)^46-49^. Similarly, the pooled prevalence rate for depression in Spain population is 35%, which is much higher than reported in Bareeqa et al. (2020) (27%), Pappa et al. (2020) (23%), Krishnamoorthy et al. 2020 (26%), and Ren et al. 2020 (28%)^46-49^.The prevalence rates in Spain exceed the reported prevalence rates of mental health issues in China, suggesting that, a higher percent of people in Spain suffered greater mental health issues. Differences across countries may be due to differences in social determinants and exposure to media ^50^.

In addition, the pooled prevalence of anxiety in Spain is higher than those in 10 counties (China, India, Japan, Iran, Iraq, Italy, Nepal, Nigeria, Spain, and UK) (32%)^51^ and is similar to those in 17 countries (i.e., China, Singapore, India, Japan, Pakistan, Vietnam, Iran, Israel, Italy, Spain, Turkey, Denmark Greece, Argentina, Brazil, Chile and Mexico) (33%)^5^. The pooled prevalence for depression in Spain is also higher than the pooled prevalence in a study which had over 17 countries reported by Luo et al. ^5^ (28%) and higher than another study of 10 countries reported by <u>Salari</u> (34%)^51^, however the prevalence rate for depression in Spain is still lower than Italy - the highest country (67%)^5^.

The pooled prevalence rate for insomnia among the Spain population (52%) is much higher than those in Germany (39%)^52^ and China (27%) as well as the pooled prevalence in the 13 countries including Australia, Bahrain, Canada, Germany, Greece, Iraq, India, Mexico and USA (36%), the general population (32%) and healthcare workers (36%) and is similar to Italy (55%) and France (51%) - two countries with the highest prevalence rate.

The pooled prevalence rate of mental illness symptoms of the student population in Spain (50%) is higher than other meta-analyses such as 28% on anxiety reported by Lasheras et al. ^53^ and 32% on anxiety, 36% on depression, and 33% on sleep disturbance reported by Deng et al. ^43^.

### 4.3 Practical Implications

The findings of this meta-analysis show that there were higher rates of anxiety, depression, and especially insomnia symptoms reported in Spain, compared to all the other countries reported apart from other nearby European countries of Italy and France. The evidence suggests the need to pay attention to the high prevalence rate of insomnia. As the literature shows that insomnia can be caused by fear of infection, quarantine, and death, making safety precautions such as masks and isolation methods can be critical ^54,55^. It is also reported that extensive use of social media during COVID-19 is associated with higher rates of anxious symptoms, distress, and insomnia related symptoms.

The evidence from this study shows that the frontline HCWs and students suffered more than the general population on anxiety, depression, and insomnia symptoms which suggests that policymakers and healthcare organizations need to prioritize frontline HCWs and students in this ongoing pandemic. Frontline HCWs are at a greater risk for mental illness symptoms which could provide the impetus for policies and strategic priorities towards building resiliency in HCWs. Protective measures, such as intercessions, self-help resources and specified care of the healthcare workers are of the utmost importance ^56^. In additional reason for prioritization is to preempt a possible rise in suicide associated with distress (e.g., mental health distress, economic insecurity) during COVID-19. ^57^

The finding that insomnia had the highest prevalence rate relative to anxiety and depression in Spain suggests a clarion call to probe populations for the presence of insomnia during the COVID-19 pandemic. It is also important to explore underlying causative factors resulting in the extraordinarily high rates. Moreover, our findings on the high rate of insomnia compared to anxiety and depression, which have been the most studied mental health issues, acts as a call for future research on insomnia, especially given that more than 90% of existing empirical studies studied anxiety and depression. ^58^

Lastly, to our surprise, no studies have examined the mental health issues of general HCWs in Spain. A possible reason for this could be that all the general HCW’s are on the frontline given the needs of the country. More research overall is needed to investigate the prevalence of HCWs, particularly general HCWs.

### 4.4 Limitations and future research

Our meta-analyses had several limitations. Among the 41 samples included in this meta-analysis, 32 samples (78.1%) investigated the prevalence of the general population in Spain with only 4 samples on frontline HCWs and 5 samples on students. The lack of sufficient data from HCWs and students limited the reliability of the pooled prevalence rates.

Secondly, we found that articles using different instruments and inconsistent cut-off scores, which makes it difficult and often impossible to accumulate and compare research findings. For example, the prevalence of anxiety measured by GAD-7 (31%) and DASS-21 (15%), the two most popular scales, differed significantly in Spain (Appendix 2). Additionally, several articles reported an “overall” prevalence without indicating the cut-off points used which makes it impossible to know which outcome level they used. We suggest future research to specify the severity levels and the cut-off points used, especially to report overall prevalence. Third, non-English articles were not included and hence could create biases in our study.

## 5 Conclusion

This is the first meta-analysis to provide evidence on the pooled prevalence rates of mental distress in Spain during the COVID-19 pandemic. Our results indicate that the people in Spain are experiencing psychological and emotional distress during COVID-19, and possibly greater than some other countries. Therefore, it is critical to identify the prevalence of specific psychological symptoms of the key populations to guide mental health assistance and resource allocation effort in the unprecedented crisis.

## Data Availability

It is available after a request.

## Credit author statement

**RZC**: *Investigation, Data curation, Visualization, Writing – original draft, Writing – review & editing*. **SXZ**: *Conceptualization, Methodology, Validation, Formal analysis, Investigation, Data curation, Visualization, Writing – original draft, Writing – review & editing, Supervision*. **WX, AY, RKD, BZC, AD, SM**: *Data curation*, **RSM**: *Writing – review & editing*, **XW:** *Investigation, Data curation*. All authors were involved in approving the manuscript. The corresponding author attests that all listed authors meet authorship criteria and that no others meeting the criteria have been omitted.

## Transparency declaration

*The corresponding author affirms that this manuscript is an honest, accurate, and transparent account of the study being reported; that no important aspects of the study have been omitted; and that any discrepancies from the study as planned (and, if relevant, registered) have been explained*.

## Ethical approval

*Not applicable*

## Patient and public involvement

*No patient or public was involved in a systematic review and meta-analysis*

---

**Table S1:** The search string used in this systematic review and meta-analysis. (Feb 6, 2021)

| Search | Search topic | Search keywords (titles, abstracts, and subject headings) with Boolean operators |
| --- | --- | --- |
| 1 | Exposure/<br>Context | 2019-nCoV OR 2019nCoV OR COVID-19 OR SARS-CoV-2 OR ((Wuhan AND coronavirus) |
| 2 | Outcome of<br>interest | "depressi*" OR "anxi*"OR "insomnia" OR "sleep symptoms" OR "sleep issue" OR "sleep problem" OR "mental illness symptoms" OR "mental issue" OR "mental problem" OR "psychiatric symptoms" OR "psychiatric issue" OR "psychiatric problem" |
| 3 | Epidemiological<br>phenomenon | “Prevalence” OR “Incidence” OR "rate*" OR "ratio*" OR “Epidemiolog*” OR “risk factor*” OR “relative risk” OR “odds ratio” OR “risk ratio” OR “disease burden” OR “Proportion” OR “percent*” |
| 4 | Language | English |
| 5 | Time | 2020 - 2021 |
|  | Overall search | 1 AND 2 AND 3 AND 4 AND 5 |

**Appendix 2.**
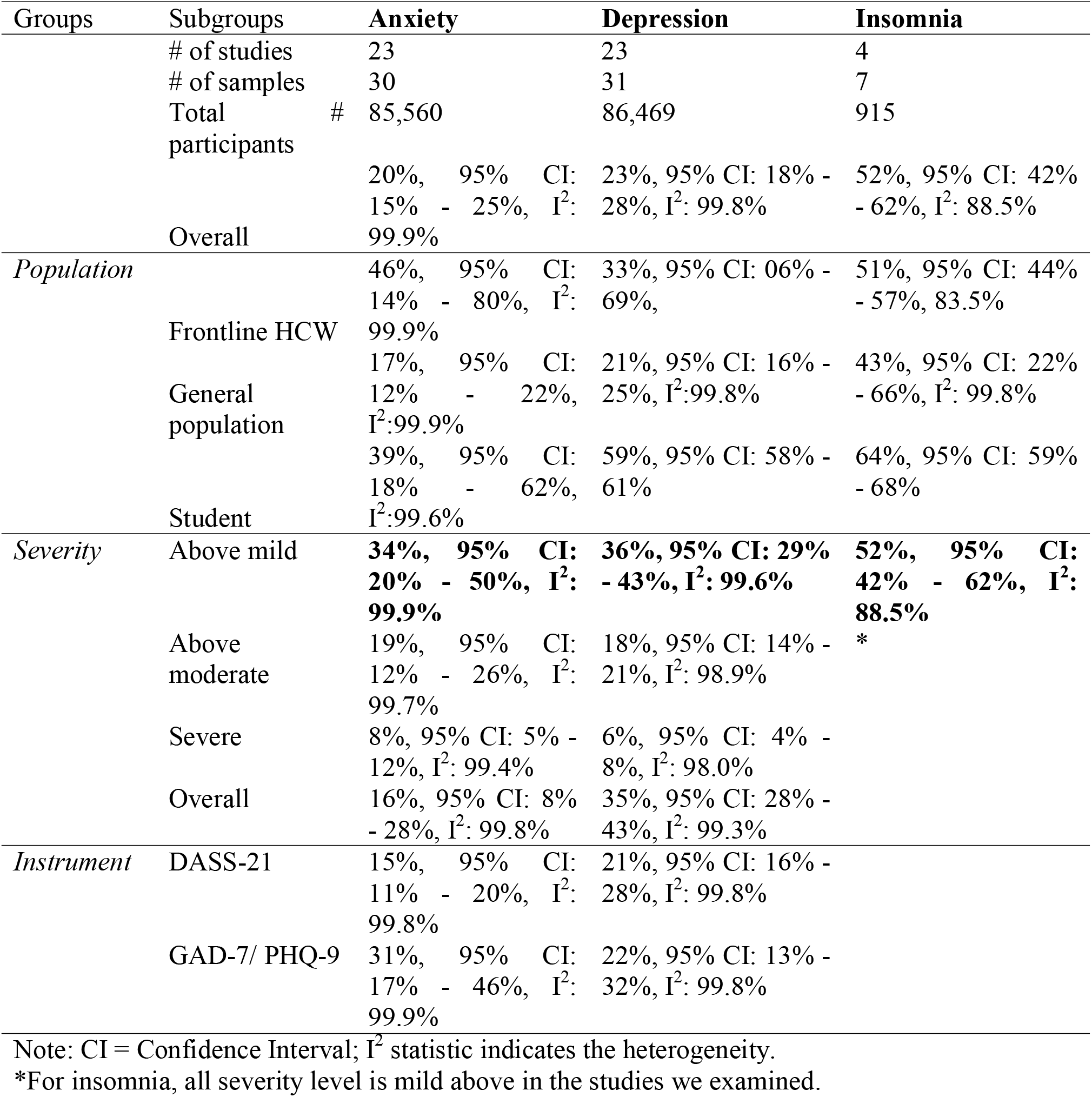
Subgroup analyses of the prevalence of anxiety, depression, and insomnia.

